# Tocilizumab shortens time on mechanical ventilation and length of hospital stay in patients with severe COVID-19: a retrospective cohort study

**DOI:** 10.1101/2020.07.29.20164160

**Authors:** Johannes Eimer, Jan Vesterbacka, Anna-Karin Svensson, Bertil Stojanovic, Charlotta Wagrell, Anders Sönnerborg, Piotr Nowak

## Abstract

**Background:** Hyperinflammation is a key feature of the pathogenesis of COVID-19 with a central role of the interleukin-6 pathway. We aimed to study the impact of the IL-6 receptor antagonist tocilizumab on the outcome of patients admitted to the intensive care unit (ICU) with acute respiratory distress syndrome (ARDS) related to COVID-19.

**Methods:** Eighty-seven patients with confirmed SARS-CoV-2 infection and moderate to severe ARDS were included (n tocilizumab = 29, n controls = 58). A matched cohort was created using a propensity score. The primary endpoint was 30-day all-cause mortality, secondary endpoints included ventilation-free days and length of stay.

**Results:** No difference was found in 30-day all-cause mortality in patients treated with tocilizumab compared to controls (17.2% vs. 32.8%, p = 0.2; HR = 0.52 [0.19 - 1.39], p = 0.19). Ventilator-free days were 19.0 (IQR 12.5 - 20.0) versus 9 (IQR 0.0 - 18.5; p = 0.04), respectively. A higher rate of freedom from mechanical ventilation at 30 days was achieved in patients receiving tocilizumab (HR 2.83 [1.48 - 5.40], p < 0.002). Median length of stay in ICU and total length of stay were reduced by 8 and 9.5 days in patients treated with tocilizumab. Similar results were obtained in the analysis of the propensity score matched cohort.

**Conclusions:** Treatment of critically ill patients with ARDS due to COVID-19 with tocilizumab was not associated with reduced 30-day all-cause mortality, but shorter duration on ventilatory support as well as shorter overall length of stay in hospital and in ICU.

---

Dear editor,

among patients with COVID-19 who require treatment in intensive care for acute respiratory distress syndrome (ARDS), mortality rates have been reported between 16 – 78% (1). In patients who are discharged alive, an increased risk of sequelae from COVID-19 is anticipated (2). The hyperinflammatory response induced by SARS-CoV-2 is pivotal in the pathogenesis of COVID-19 and is accompanied by an upregulated expression of interleukin 6 (IL-6) that correlates with disease severity (3). Tocilizumab, a monoclonal antibody against the IL-6 receptor originally licensed for the use in rheumatoid arthritis, is also approved for treatment of chimeric antigen receptor T cell–related cytokine release syndromes and secondary hemophagocytic syndromes that share important features with the hyperinflammatory phase in COVID-19. Several small studies from China and Europe have reported promising results of the treatment with tocilizumab in patients with COVID-19, preventing the need for admission to an intensive care unit and improving clinical outcomes (4,5). We aimed to evaluate the impact of treatment with tocilizumab compared to routine care on important clinical outcomes in critically ill patients admitted to an intensive care unit with ARDS due to COVID-19.

## Methods

We conducted a retrospective cohort study at Karolinska University Hospital Huddinge between 11th March and 15th April 2020 (regional ethical approval: Drn 2020-3139). Patients over 18 years with confirmed SARS-CoV-2 infection were eligible when admitted to the intensive care unit (ICU) for severe ARDS and were followed for 30 days from admission to ICU until discharge from hospital or until death, whichever occurred first.

All patients who received tocilizumab before admission to or on ICU during the study period were included. Consideration of treatment with a single dose of tocilizumab at 8 mg/kg was at the discretion of the attending physician and required consultation of at least two members of an expert panel of infectious disease specialists as well as the fulfilment of specific criteria based on respiratory and inflammatory parameters. The control group consisted of consecutively admitted patients to the same ICU receiving routine care only (see supplement for details). The primary outcome was 30-day all cause mortality after admission to ICU (= day 0). Secondary outcomes were time to freedom from mechanical ventilation, number of ventilator-free days in survivors, length of stay in ICU and in hospital. Clinical endpoints were assessed in the native cohort and in a sub-cohort of patients matched by a propensity score (see supplement for detailed methodology).

## Results

Of 87 patients in the cohort, 29 received tocilizumab and 58 patients received routine care only (control group). Twenty-two patients (n=22) from each group were matched within a propensity-score matched sub-cohort. Notable differences between groups in the native cohort included a higher proportion of male patients in the tocilizumab group and a lower body mass index. Respiratory parameters were comparable upon admission to the hospital and upon admission to ICU. In accordance with the prespecified treatment criteria, inflammatory biomarkers were higher in the tocilizumab group upon admission to ICU. Baseline comparability was improved in the propensity score matched sub-cohort (Suppl. table 1). As to the outcomes, the difference in all cause mortality at 30 days was not statistically significant (HR = 0.52, 95% CI 0.19 - 1.39, p = 0.19) (Figure 1). However, patients receiving tocilizumab had significantly more ventilator-free days (Suppl. table 2). Freedom from mechanical ventilation was achieved earlier and in a higher proportion of patients (HR 2.83, 95% CI = 1.48 - 5.40, p = 0.002) (Figure 1). Length of stay in ICU and length of stay in hospital were both significantly shorter in patients treated with tocilizumab (Figure 1). The rate of serious secondary bacterial infections upon treatment with tocilizumab was comparable to controls. No serious adverse events attributable to the intervention were recorded. Analysis of the matched sub-cohort revealed consistent results across all outcomes (Figure 1, Suppl. table 2).

**Figure 1:**
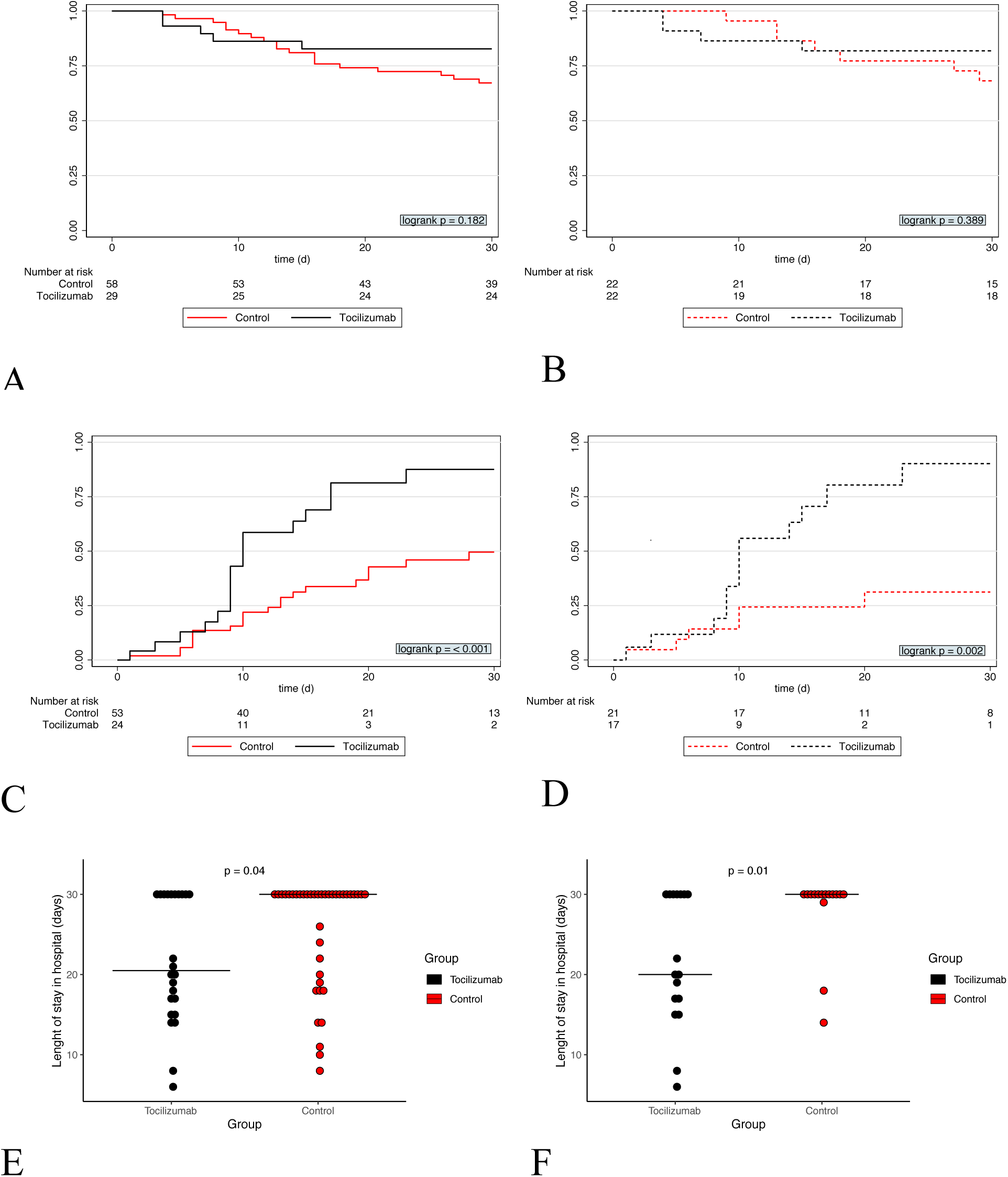
Upper panel: Cumulative 30-day survival rate depicted as Kaplan-Meier plots in the native (A) and propensity score matched cohort (B). Central panel: Cumulative rate of freedom from mechanical ventilation among invasively ventilated patients depicted as Kaplan-Meier plots in the native (C) and propensity score matched cohort (D). Lower panel: Total length of hospital stay, calculated from admission to the hospital until discharged alive or alive and hospitalized at 30 days (i.e. end of follow-up) in the (E) native cohort and (F) propensity score matched cohort. The horizontal thin line represents the median.

## Discussion

In this retrospective cohort study, the administration of tocilizumab did not reduce all cause mortality but was associated with a shorter time on mechanical ventilation and a shorter length of stay in hospital and in ICU in critically ill patients with ARDS due to COVID-19. The treatment was well tolerated and not associated with an increased rate of serious adverse events during the study period. Results were confirmed in a propensity-score matched sub-cohort.

Mortality in our study was low compared to previous reports (1). This may be explained by a comparatively low prevalence of comorbidities and a low median age of 56 years [IQR 49-64]. Four out of five (4/5, 80%) patients who died in ICU following treatment with tocilizumab died within nine days from admission to the hospital from multiple organ failure. Deaths occurred earlier than in the control group (median 8 vs. 14 days). All of these individuals presented with significant comorbidities. We hypothesize that the early deaths among patients in the intervention group represent a proportion of patients with a poor baseline prognosis and a potentially irreversible hyperinflammatory syndrome. Patients receiving tocilizumab had significantly more ventilator-free days compared to controls and achieved freedom from mechanical ventilation earlier. A pronounced divergence in respiratory recovery between groups was observed after day 10 (Figure 1). This may represent a lag time of clinical improvement following the rapid onset of action of tocilizumab. Generally, time on the ventilator correlates with subsequent complications such as infections, cognitive impairment and critical illness neuro-myopathy (6). Thus, our findings suggest a potential role of tocilizumab in the prevention of post-ICU sequelae from severe COVID-19. If confirmed in a prospective randomized trial, the substantial reduction in length of stay in ICU observed in our cohort would most likely render the intervention highly cost-efficient, with a single dose of tocilizumab (8mg/kg, adult patient of 75 kg) being priced around 3035.93 $ in the US according to recently published model (7).

We acknowledge several limitations. Our study was designed as a retrospective cohort study at a single academic medical center with inherent limitations to generalizability of findings and potential biases. Furthermore, the limited number of patients treated with tocilizumab restricted the power to detect a significant 30-day mortality difference. A strength of the study is the 30-day follow up exceeding previous reports on immunomodulatory treatment of COVID-19 and adding further evidence to the course of disease in critically ill patients with COVID-19. In addition to that, the analysis after propensity score-based matching did not significantly alter the results, thus reducing the likelihood of measured confounders being the sole explanation of the differences in outcomes.

In summary, our findings indicate that treatment with tocilizumab of critically ill patients with severe ARDS due to COVID-19 may reduce time on mechanical ventilation and overall length of stay in ICU and in hospital. Treatment appears to be safe. Data from randomized controlled trials are needed to confirm the results and establish causality.

## Data Availability

Data are availible on request from other scientists

## Acknowledgments

None.

## Bibliography

1. Grasselli G, Zangrillo A, Zanella A, Antonelli M, Cabrini L, Castelli A, et al. Baseline Characteristics and Outcomes of 1591 Patients Infected With SARS-CoV-2 Admitted to ICUs of the Lombardy Region, Italy. JAMA. 2020 Apr 28;323(16):1574–81.

2. Mo X, Jian W, Su Z, Chen M, Peng H, Peng P, et al. Abnormal pulmonary function in COVID-19 patients at time of hospital discharge. European Respiratory Journal [Internet]. 2020 Jan 1 [cited 2020 Jun 5]; Available from: https://erj.ersjournals.com/content/early/2020/05/07/13993003.01217-2020

3. Blanco-Melo D, Nilsson-Payant BE, Liu W-C, Uhl S, Hoagland D, Møller R, et al. Imbalanced Host Response to SARS-CoV-2 Drives Development of COVID-19. Cell. 2020 May 28;181(5):1036-1045.e9.

4. Capra R, De Rossi N, Mattioli F, Romanelli G, Scarpazza C, Sormani MP, et al. Impact of low dose tocilizumab on mortality rate in patients with COVID-19 related pneumonia. Eur J Intern Med. 2020 Jun;76:31–5.

5. Xu X, Han M, Li T, Sun W, Wang D, Fu B, et al. Effective treatment of severe COVID-19 patients with tocilizumab. Proceedings of the National Academy of Sciences. 2020 Apr 29;202005615.

6. Desai SV, Law TJ, Needham DM. Long-term complications of critical care. Crit Care Med. 2011 Feb;39(2):371–9.

7. Schmier J, Ogden K, Nickman N, Halpern MT, Cifaldi M, Ganguli A, et al. Costs of Providing Infusion Therapy for Rheumatoid Arthritis in a Hospital-based Infusion Center Setting. Clinical Therapeutics. 2017 Aug 1;39(8):1600–17.

